# Incidence of dementia after a recent cancer diagnosis among people with HIV

**DOI:** 10.64898/2026.02.12.26346206

**Authors:** Corinne E. Joshu, Maylin Palatino, Jacqueline E. Rudolph, Karine Yenokyan, Keri Calkins, Xiaoqiang Xu, Yiyi Zhou, Eryka Saylor, Bryan Lau

## Abstract

**Objective:** To evaluate risk of dementia after cancer diagnosis among Medicaid beneficiaries with HIV.

**Design:** Longitudinal observational study of Medicaid enrollment, inpatient, and outpatient claims data from 14 states, 2001-2015.

**Methods:** Beneficiaries aged 18-64 with HIV and ≥6 months of enrollment were matched 1:1 on cancer status by age, sex, race, year, and state. We estimated the weighted cumulative incidence functions (CIFs) of dementia at 1, 2, and 5 years after cancer diagnosis using the Aalen-Johansen estimator to account for the competing risk of death and cluster stratified analyses to account for matching. We calculated the corresponding risk differences (RD) and 95% confidence intervals (CI) using nonparametric bootstrap.

**Results:** At 5 years, the CIF of dementia was 9.6% (95%CI: 8.2, 11.6) and 4.7% (95%CI: 3.7, 6.1) among those with and without AIDS-defining cancer, respectively (RD: 4.9%; 95%CI: 2.9, 7.0). At 5 years, the CIF of dementia was 7.1% (95%CI: 5.9, 7.8) and 5.3% (95%CI: 4.2, 6.2) among those with and without non-AIDS-defining cancer, respectively (RD: 1.8%; 95%CI: 0.34, 2.9). Dementia incidence appeared higher among beneficiaries with lung cancer (2yr RD: 1.9%; 95%CI: 0.01, 5.2) and beneficiaries ≤50 with colon cancer (2yr RD: 4%; 95%CI: 0.3, 10.5), but lower among beneficiaries ≤50 with prostate cancer (2yr RD: −1.9%; 95%CI: −2.3, −1.6). Dementia incidence did not differ among beneficiaries with and without breast cancer.

**Conclusions:** Dementia risk may be increased among people with HIV with certain cancers, including AIDS-defining cancers. Dementia risk appears to vary by cancer type and age at diagnosis.

## Introduction

People with HIV (PWH) have an elevated risk of cognitive impairment ^[1-3]^. Effective antiretroviral therapy (ART) has led to improved life expectancy and a shift in the types of neurocognitive disorders among PWH. Prior to ART, HIV-associated neurocognitive disorders (HAND), which encompass a range of cognitive complications that can span from asymptomatic impairment identified upon targeted testing to severe impairment that impacts daily living, were common particularly among individuals with advanced HIV ^[1, 3, 4]^. Post ART, HIV-associated dementia is uncommon among PWH with well-controlled HIV ^[1, 3]^. As improvements in survival among PWH continue, there has been increased focus on better characterizing the burden of and risk factors for age-associated neurodegenerative diseases including dementias such as Alzheimer’s disease and vascular dementia ^[3, 4]^. A recent evaluation of a three large healthcare systems reported that although new cases of dementia decreased over time, PWH still had a substantially higher incidence and prevalence of dementia than those without HIV ^[3]^. Among PWH, older age, lower socioeconomic status, and diagnosis of AIDS have been associated with an increased risk of dementia ^[4]^. More work is needed to better characterize risk factors for dementia among PWH.

Cancer is a significant comorbidity for people with HIV (PWH). Effective ART has also contributed to a shift in the distribution of cancer types among PWH ^[5]^. Cancers associated with immunosuppression and/or co-infection, including non-Hodgkin lymphoma and Kaposi sarcoma, have decreased while cancers common in the general population, including prostate and lung, have increased ^[5, 6]^. In the general population, prior epidemiologic studies have consistently reported an inverse association between cancer and dementia, which is paradoxical given shared risk factors, treatment effects, and psychological stress experienced by those with a history of cancer ^[7, 8]^. Meta-analyses of population-based studies have reported modest but consistent inverse associations between cancer and dementia, but have highlighted the challenges in accounting for biases, including competing risks and survival bias, in those findings ^[7, 8]^. To our knowledge, the impact of a recent cancer diagnosis on dementia incidence among PWH has not been well-characterized.

Thus, we evaluated the incidence of dementia among PWH with and without incident cancer who were enrolled in Medicaid between 2000 and 2015 in one of 14 US states. Briefly, Medicaid is a joint federal and state program that provides health coverage to individuals that meet certain criteria including those who are low-income; 40% of PWH in the US are covered by Medicaid ^[9]^. To address concerns of prior study designs among people without HIV, we matched PWH with cancer to PWH without cancer and estimated dementia incidence at 1, 2, and 5 years post-cancer diagnosis while accounting for the competing risk of death.

## Methods

### Study Sample

We utilized the enrollment, inpatient, and outpatient claims data from the Centers for Medicare and Medicaid Services (CMS) beneficiaries between 2001 and 2015 in one of 14 states: Alabama, California, Colorado, Florida, Georgia, Illinois, Maryland, Massachusetts, New York, North Carolina, Ohio, Pennsylvania, Texas, and Washington. To ensure complete capture of medical claims and equivalent opportunity for services, we restricted the analysis to beneficiaries 18-64 years old with HIV, full benefits, no dual enrollment in Medicare or private insurance, and ≥6 months of continuous enrollment. HIV was defined as having one inpatient claim or two outpatient claims with an HIV-related ICD-9 code within 2 years; the first service date was used at date of diagnosis (Table S1).^[10]^ Because incident, not prevalent, cancer was our exposure of interest, beneficiaries with any evidence of a cancer diagnosis on any claim within the first six months of enrollment were excluded (Table S1).^[10]^ The Johns Hopkins Bloomberg School of Public Health Institutional Review Board determined that this secondary analysis of existing Medicaid claims data meets the criteria for exemption.

### Exposure

We defined incident cancer as having one inpatient claim or two outpatient claims with a cancer-related ICD-9 code within 2 years; the first service date was used as date of diagnosis (Table S1) after the first 6 months of continuous enrollment.^[10]^ We identified AIDS-defining cancers (hereafter ADCs; Kaposi’s sarcoma, non-Hodgkin lymphoma, or cervical cancer), non-AIDS defining cancers (hereafter NADCs; all non-ADC cancers), and lung, colon, breast, and prostate cancers.

### Outcome

We defined incident dementia as having one inpatient claim or two outpatient claims with a dementia-related ICD-9 code within 2 years; the first service date was used as date of diagnosis (ICD-9 codes in Table S1) ^[10]^. Because we sought to evaluate incident dementia diagnoses after a cancer diagnosis, beneficiaries with any evidence of a dementia diagnosis on any claim prior to the cancer diagnosis/index date for the matched cluster were excluded (see Matching).

### Covariates

We obtained demographic and enrollment information from the personal summary file, including sex, race (non-Hispanic White, non-Hispanic Black, Hispanic, other), US state, and dates of birth, death, enrollment, and disenrollment. We defined Charlson comorbidity conditions -- myocardial infarction (MI), congestive heart failure (CHF), peripheral vascular disease (PVD), cerebrovascular disease (CVD), chronic pulmonary disease (COPD), rheumatic disease, peptic ulcer disease, liver disease, hemiplegia or paraplegia, renal disease, and diabetes--as having one ICD-9 diagnosis code on any claim prior to the cancer diagnosis/index date for the matched cluster (Table S1).^[11]^ Using the prescription data file, we calculated the monthly ART medication possession ratio (MPR) as the proportion of days per month with ART supply using the ART prescription fill date and number of days’ supply. We categorized beneficiaries as having any ART use if MPR>0 for any month prior to the cancer diagnosis/index date for the matched cluster and as being ART adherent if the mean monthly MPR was greater than 80% in the 6 months prior the cancer diagnosis/index date for the matched cluster.

### Matching

We matched beneficiaries by cancer status for incident diagnoses of ADCs, NADCs and lung, breast, colon, and prostate cancers (Table 1; Supplemental methods). Briefly, we used incidence density matching with replacement to match beneficiaries with cancer to beneficiaries without cancer at the date of cancer diagnosis. Date of cancer diagnosis was set as the index date and analytic baseline for each matched cluster. Beneficiaries were matched (1:1 or 1:many, table 1) by age within 2 years, sex, race, US state, and enrollment year within 2 years. To better account for shared risk factors, we conducted sensitivity analyses: (1) lung cancer cases were matched to cancer-free beneficiaries with COPD to better account for smoking, and (2) colon cancer cases were matched to cancer-free beneficiaries with any Charlson comorbidity to better account for differences in metabolic and cardiovascular health.

**Table 1a.** Demographic characteristics, dementia incidence and median follow-up time of matched Medicaid beneficiaries with HIV by cancer type, 2001-2015.

| Characteristic | ADC<sup>1</sup> |  | NADC<sup>2</sup> |  | Lung cancer |  |
| --- | --- | --- | --- | --- | --- | --- |
|  | Without | With | Without | With | Without | With |
| N<sup>3</sup> | 4288 | 4288 | 8883 | 8883 | 91,421 | 1774 |
| Mating Ratio | 1:1 |  | 1:1 |  | 93:1 |  |
| Age at baseline, Median [IQR]<sup>4</sup> | 44.4<br>[38.2, 50.6] | 44.4<br>[38.5, 50.9] | 50.92<br>[44.56, 56.64] | 51.26<br>[44.89, 56.90] | 52.38<br>[47.24, 57.54] | 52.82<br>[47.54, 57.82] |
| Sex, N (%) |  |  |  |  |  |  |
| Female | 1734 (40.4) | 1734 (40.4) | 3716 (41.8) | 3716 (41.8) | 745 (42.0) | 745 (42.0) |
| Male | 2554 (59.6) | 2554 (59.6) | 5167 (58.2) | 5167 (58.2) | 1029 (58.0) | 1029 (58.0) |
| Race/Ethnicity, N (%) |  |  |  |  |  |  |
| Non-Hispanic White | 930 (21.7) | 930 (21.7) | 1927 (21.7) | 1927 (21.7) | 407 (22.9) | 407 (22.9) |
| Non-Hispanic Black | 2266 (52.8) | 2266 (52.8) | 4893 (55.1) | 4893 (55.1) | 1051 (59.2) | 1051 (59.2) |
| Hispanic | 447 (10.4) | 447 (10.4) | 722 (8.1) | 722 (8.1) | 87 (4.9) | 87 (4.9) |
| Other | 645 (15.0) | 645 (15.0) | 1341 (15.1) | 1341 (15.1) | 229 (12.9) | 229 (12.9) |
| US State, N (%) |  |  |  |  |  |  |
| AL | 55 (1.3) | 55 (1.3) | 134 (1.5) | 134 (1.5) | 21 (1.2) | 21 (1.2) |
| CA | 625 (14.6) | 625 (14.6) | 977 (11.0) | 977 (11.0) | 166 (9.4) | 166 (9.4) |
| CO | 17 (0.4) | 17 (0.4) | 18 (0.2) | 18 (0.2) | NR<sup>4</sup> | NR<sup>4</sup> |
| FL | 699 (16.3) | 699 (16.3) | 1349 (15.2) | 1349 (15.2) | 248 (14.0) | 248 (14.0) |
| GA | 198 (4.6) | 198 (4.6) | 413 (4.6) | 413 (4.6) | 76 (4.3) | 76 (4.3) |
| IL | 254 (5.9) | 254 (5.9) | 588 (6.6) | 588 (6.6) | 120 (6.8) | 120 (6.8) |
| MA | 111 (2.6) | 111 (2.6) | 240 (2.7) | 240 (2.7) | 59 (3.3) | 59 (3.3) |
| MD | 201 (4.7) | 201 (4.7) | 420 (4.7) | 420 (4.7) | 118 (6.7) | 118 (6.7) |
| NC | 137 (3.2) | 137 (3.2) | 295 (3.3) | 295 (3.3) | 74 (4.2) | 74 (4.2) |
| NY | 1548 (36.1) | 1548 (36.1) | 3625 (40.8) | 3625 (40.8) | 721 (40.6) | 721 (40.6) |
| OH | 83 (1.9) | 83 (1.9) | 137 (1.5) | 137 (1.5) | 29 (1.6) | 29 (1.6) |
| PA | 91 (2.1) | 91 (2.1) | 193 (2.2) | 193 (2.2) | 49 (2.8) | 49 (2.8) |
| TX | 242 (5.6) | 242 (5.6) | 454 (5.1) | 454 (5.1) | 85 (4.8) | 85 (4.8) |
| WA | 27 (0.6) | 27 (0.6) | 40 (0.5) | 40 (0.5) | NR<sup>5</sup> | NR<sup>5</sup> |
| Enrollment period, N (%) |  |  |  |  |  |  |
| 2001-2005 | 1811 (42.2) | 1811 (42.2) | 2776 (31.3) | 2776 (31.3) | 585 (33.0) | 585 (33.0) |
| 2006-2010 | 1372 (32.0) | 1372 (32.0) | 3013 (33.9) | 3013 (33.9) | 593 (33.4) | 593 (33.4) |
| 2011-2015 | 1105 (25.8) | 1105 (25.8) | 3094 (34.8) | 3094 (34.8) | 596 (33.6) | 596 (33.6) |
| Incident dementia, N (%) | 134 (3.1) | 265 (6.2) | 302 (3.4) | 389 (4.4) | 3,398 (3.7) | 57 (3.2) |
| Follow-up years, Median [IQR]<sup>4</sup> | 3.0<br>[1.2, 5.0] | 1.1<br>[0.3, 3.3] | 2.7<br>[1.1, 5.0] | 1.2<br>[0.4, 3.2] | 3.1<br>[2.0, 3.6] | 0.6<br>[0.2, 1.4] |

<sup>1</sup>ADC: AIDS-defining cancer <sup>2</sup>Non-AIDS defining cancer <sup>3</sup>The number of beneficiaries without cancer and unique beneficiaries that were matched to at least 1 cancer case. Beneficiaries were matched using incidence density sampling with replacement. <sup>4</sup>IQR: Interquartile range <sup>5</sup>NR: Cell sizes below <11 are not reportable.

**Table 1b.**
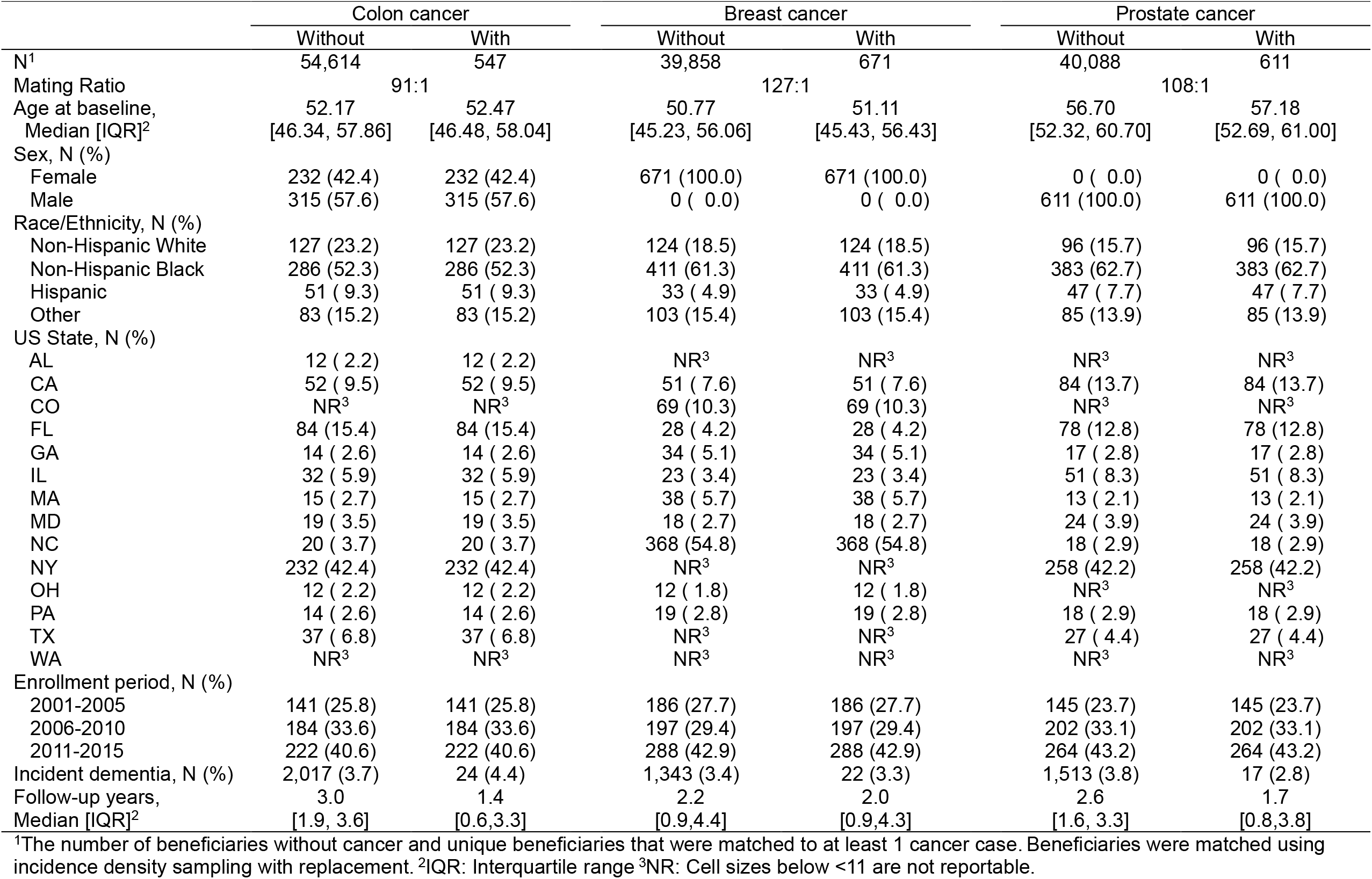
Demographic characteristics, dementia incidence and median follow-up time of matched Medicaid beneficiaries with HIV by cancer type, 2001-2015.

### Statistical Analysis

We calculated means and proportions of baseline characteristics of beneficiaries and their matches for each cancer category. Beneficiaries were followed from the date of analytic baseline until dementia diagnosis, death, 65^th^ birthday, disenrollment, or 12/31/15, whichever came first. We estimated the crude and weighted cumulative incidence functions (CIFs) of dementia among those with and without cancer using the Aalen-Johansen estimator to account for the competing risk of death, employing robust variance estimation to account for matching. Weighted CIFs were estimated using inverse probability weighting; exposure (cancer) weights and informative censoring weights were estimated and combined (Supplemental Methods). We calculated the crude and weighted risks of dementia and death at 1, 2, and 5 years after analytic baseline for beneficiaries with and without cancer and the corresponding risk differences (RD). The 95% confidence intervals (CI) of the risks and RDs were estimated using the 2.5^th^ and 97.5^th^ percentile of the point estimates from 500 bootstrap resamples. Analyses were also stratified by cancer type and age at index date (18-49; 50-64 years). Risk of dementia and RD estimates accounting for ART adherence are described in main text; crude estimates, estimates accounting for any ART, and estimates for death are provided in the supplemental tables/figures.

## Results

Demographic characteristics between beneficiaries with and without cancer were similar due to matching (Table 1). Across cancer types, beneficiaries with ADCs were younger than beneficiaries with other cancer types and more likely to be diagnosed in the earliest observation period (2001-2005).

### AIDS-Defining Cancers (ADCs)

The 1-year risk of dementia was 4.7% (95%CI: 3.9,5.4) among those with an ADC as compared to 1.1% (95%CI: 0.8,2.4) among those without an ADC (Figure 1, Table S3). Findings were similar at 2 years post diagnosis. The 5-year risk of dementia rose to 9.6% (95%CI: 8.2,11.6) and 4.7% (95% CI: 3.7,6.1) among those with and without an ADC, respectively. Findings were similar among beneficiaries diagnosed with an ADC below age 50 and those diagnosed between 50 and 64 years. Beneficiaries with an ADC had a 4.9% (95%CI: 2.9,7.0) higher risk of dementia within 5 years of cancer diagnosis compared to those without cancer (Figure 2, Table S3). Risk of death was also higher among those with an ADC as compared to those without an ADC at 1 year (RD 14.6; 95%CI: 12.2,16.0), 2 years (RD 17.7; 95%CI: 15.8,19.3), and 5 years (RD 20.7; 95%CI: 18.3,23.4) post-diagnosis (Table S4).

**Figure 1.**
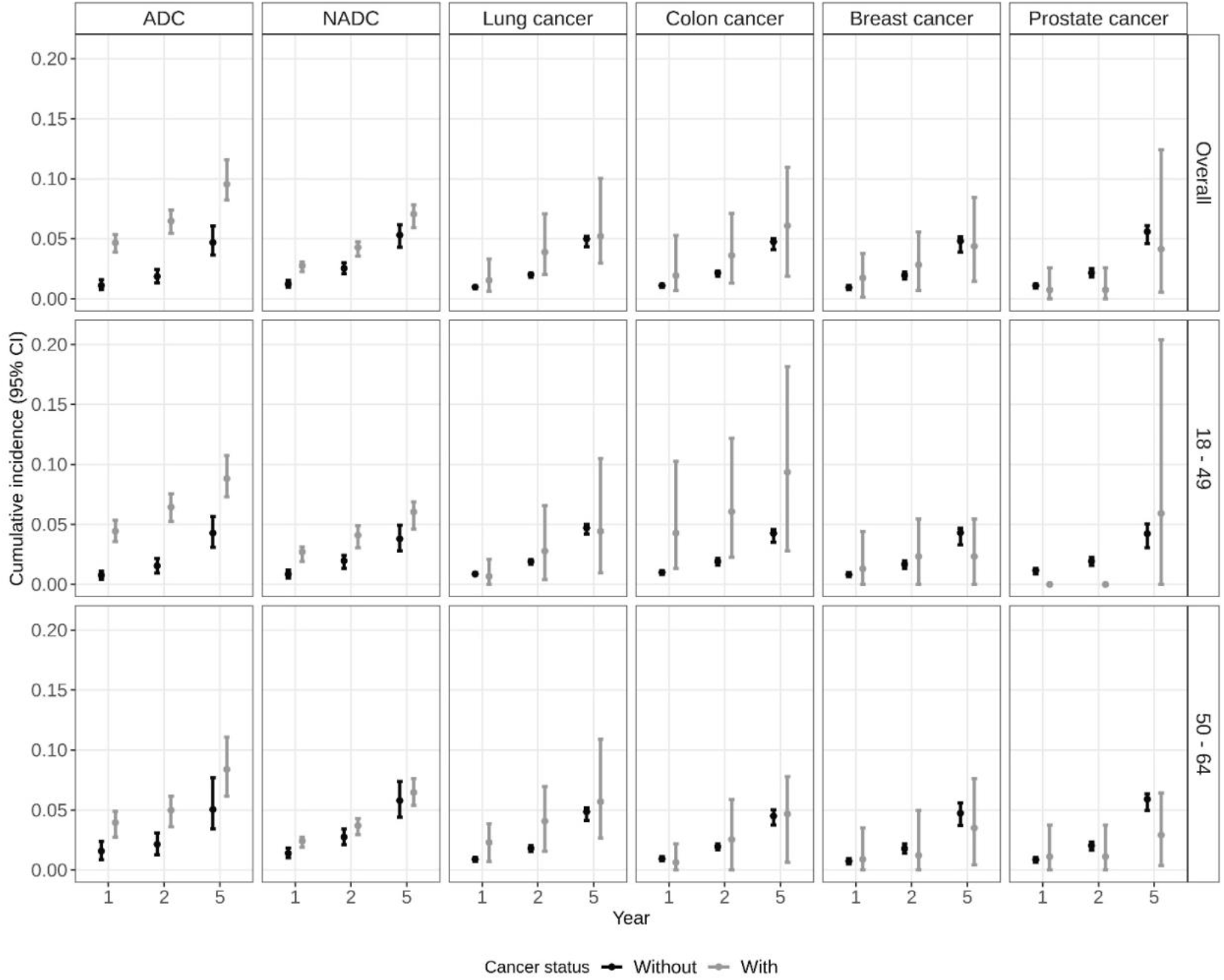
Cumulative incidence (risk%) of dementia at years 1, 2, and 5 after index date (cancer diagnosis date) among Medicaid beneficiaries with HIV matched on cancer status by age, sex, race, year, and state, overall and stratified by age. ADC: AIDS defining cancer; NADC: non-AIDS defining cancer.

**Figure 2.**
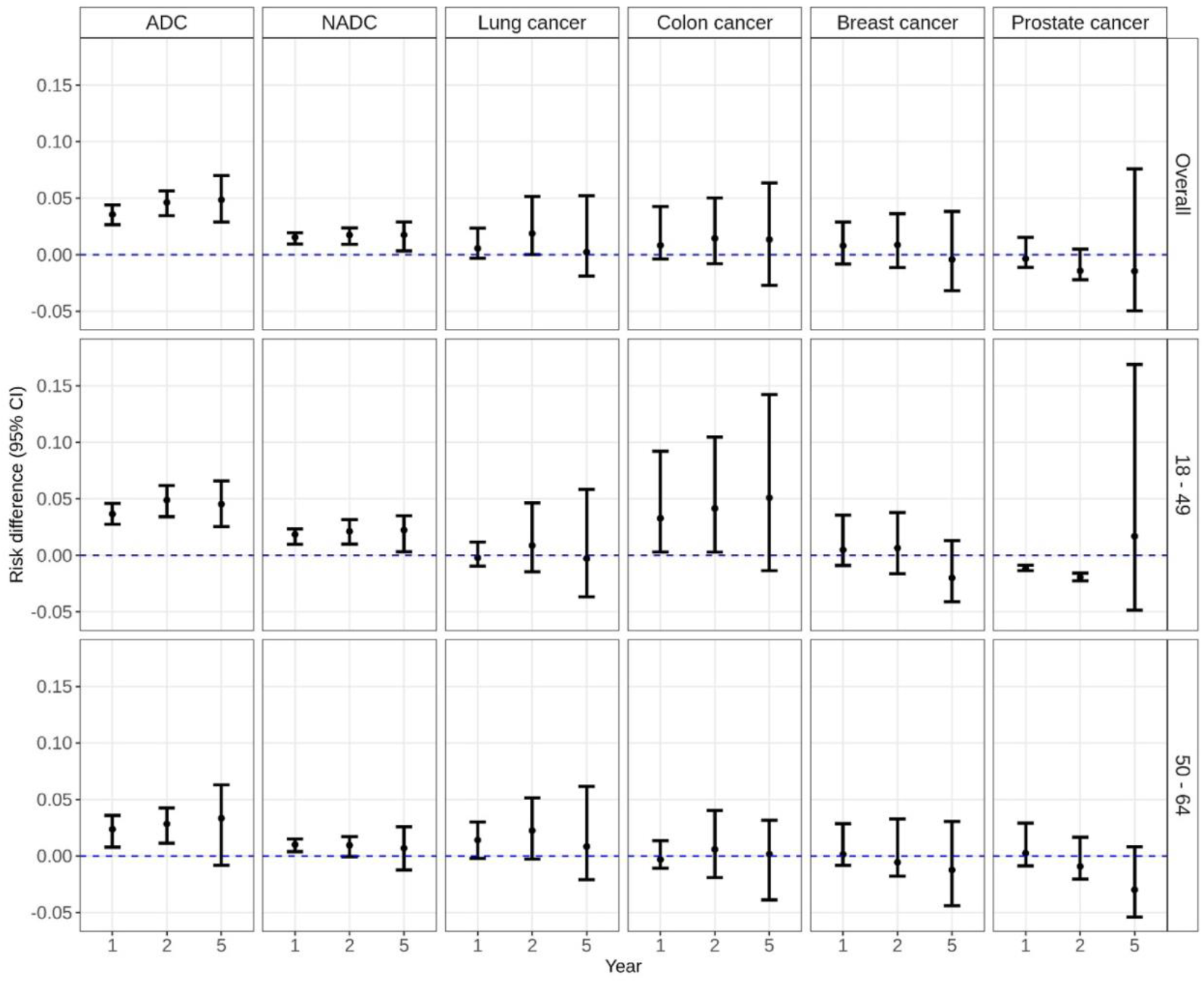
Risk difference of dementia at years 1, 2, and 5 after index date (cancer diagnosis date) among Medicaid beneficiaries with HIV matched on cancer status by age, sex, race, year, and state, overall and stratified by age. ADC: AIDS defining cancer; NADC: non-AIDS defining cancer.

### Non-AIDS Defining Cancers (NADCs)

The 1-year risk of dementia was 2.8% (95%CI: 2.2,3.1) among those with an NADC as compared to 1.2% (95%CI: 9.6,1.5) among those without an NADC (Figure 1, Table S3). Findings were similar at 2 years post diagnosis. The 5-year risk of dementia rose to 7.1% (95%CI: 5.9,7.8) and 5.3% (95%CI: 4.2,6.2) among those with and without an NADC, respectively. Beneficiaries with an NADC had a 1.8% (95%CI: 0.34,2.9) higher risk of dementia within 5 years of cancer diagnosis compared to those without cancer (Figure 2, Table S3). Among beneficiaries aged 50-64, this difference was attenuated largely due to a higher dementia incidence among beneficiaries without an NADC (Table S3). Risk of death was also higher among those with an NADC as compared to those without an NADC at 1 year (RD 14.4; 95%CI: 13.4,15.3), 2 years (RD 19.3; 95%CI:17.9,20.3), and 5 years (RD 23.6; 95%CI: 21.9,25.9) post-diagnosis (Table S4).

### Lung Cancer

The 1-year risk of dementia was 1.5% (95%CI: 0.63,3.3) among those with lung cancer and 1.0% (95%CI: 0.86,1.1) among those without lung cancer (Figure 1, Table S3). At 2-years, there was a 1.9% (95%CI: 0.01,5.2) higher risk of dementia among beneficiaries with lung cancer as compared to those without lung cancer; findings were similar but not statistically significant among beneficiaries 50-64 (2-year RD: 2.3%, 95%CI: −0.26,5.2). The 5-year risk rose to 5.2% (95%CI: 3.0,10.0) and 5.0% (95%CI: 4.3,5.2) among those with and without lung cancer, respectively. There was no difference in 5-year risk of dementia overall or when stratified by age (Figure 2). In the sensitivity analysis restricting controls to those with chronic pulmonary disorder, there was no difference in risk of dementia (Table S3). Notably, risk of death was substantially higher among those with lung cancer as compared to those without at 1 year (RD 35.1; 95%CI: 30.1,40.2) and 2 years (RD 42.9; 95%CI: 37.3,50.0). The 5-year risk of death was 60.4% (95%CI: 51.3,69.0) among those with lung cancer and compared to 6.4% (95%CI: 5.3, 7.0) among those without lung cancer (RD 54.1; 95%CI: 45.3,62.8; Table S4).

### Colon Cancer

The 1-year risk of dementia was 1.9% (95%CI: 0.68,5.3) among those with colon cancer compared to 1.0% (95%CI: 0.95,1.2) among those without colon cancer (Figure 1, Table S3). The 5-year risk rose to 6.1% (95%CI:1.9,10.9) and 4.8% (95%CI: 4.1,5.0) among those with and without colon cancer, respectively. There was no significant difference in dementia risk overall or among beneficiaries 50-64 (Figure 2, Table S3). In contrast, among those 18-50, the 1-year risk of dementia was 4.3% (95%CI: 1.3,10.2) among those with colon cancer as compared to 1.0% (95%CI: 0.8,1.1) among those without colon cancer. The 5-year risk rose to 9.4% (95%CI: 2.8,18.1) and 4.3% (95%CI: 3.5,4.6) among those with and without colon cancer, respectively. Younger beneficiaries had 4% higher incidence of dementia than their cancer-free counterparts within 2 years of diagnosis (95%CI: 0.3,10.5); the risk difference was similar but no longer statistically significant at 5 years (Figure 2, Table S3). Risk differences were similar when beneficiaries with colon cancer were matched to controls with at least one comorbid condition (Table S3). Risk of death was higher among those with colon cancer as compared to those without colon cancer at 1 year (RD 9.9; 95%CI: 6.9,15.5), 2 years (RD 15.8; 95%CI: 11.8,22.4), and 5 years (RD 24.3; 95%CI: 17.0,33.8) post-diagnosis (Table S4).

### Breast Cancer

The 1-year risk of dementia was 1.7% (95%CI: 0.2,3.8) among those with breast cancer compared to 1.0% (95%CI: 0.8,1.1) among those without breast cancer (Figure 1, Table S3). The 5-year risk rose to 4.4% (95%CI: 1.4,8.4) and 4.8% (95%CI: 3.9,5.1) among those with and without breast cancer, respectively. There was no significant difference in risk of dementia within 5 years of breast cancer diagnosis overall and when stratified by age (Figure 2, Table S3). While the risk of death was higher among those with breast cancer as compared to those without breast cancer at 1 year (RD 6.9; 95%CI: 3.3, 11.9), 2 years (RD 10.4; 95%CI: 5.7,15.0), and 5 years (RD 10.7; 95%CI: 4.6,19.8) post-diagnosis, these differences were not as large as the differences for the other cancer types evaluated (Table S4).

### Prostate Cancer

The 1-year risk of dementia was 0.7% (95%CI: 0.0,2.6) among those with prostate cancer compared to 1.1% (95%CI: 0.9,1.2) among those without prostate cancer (Figure 1, Table S3). The 5-year risk rose to 4.1% (95%CI: 0.5,12.4) and 5.6% (95%CI: 4.6,6.1) among those with and without prostate cancer, respectively. There was no significant difference in dementia risk overall or among beneficiaries 50-64 (Figure 2, Table S3). In contrast, among those 18-50, 1-year risk of dementia was 0.0% (95%CI: 0.0,0.0) among those with prostate cancer as compared to 1.2% (95%CI: 0.9,1.3) among those without prostate cancer; the 5-year risk rose to 5.9% (95%CI: 0.0,20.4) and 4.2% (95%CI: 3.1,5.1) among those with and without prostate cancer, respectively. Younger beneficiaries had 1.9% (95%CI: −2.3, −1.6) lower incidence of dementia than their cancer-free counterparts within 2 years of diagnosis, though this difference was attenuated at 5 years. In contrast to the other cancers evaluated, there was no difference in risk of death at 1- or 5-years post diagnosis; the risk of death was modestly higher among those with prostate cancer as compared to those without prostate cancer at 2 years (RD 2.8; 95%CI: 0.0, 0.7) (Table S4).

## Discussion

In this matched study of Medicaid beneficiaries with HIV, we found an increased risk of dementia after both AIDS-defining (ADC) and non-AIDS defining cancer diagnoses (NADC). Within 5 years of cancer diagnosis, dementia risk was just under 10% for PWH with an ADC and approximately 7% for PWH with an NADC as compared to approximately 5% for their cancer-free counterparts. These findings were consistent when stratified by age at cancer diagnosis. Risk differences were attenuated among those over 50 years old likely due at least in part to the increased risk of dementia in PWH without cancer among older beneficiaries. In cancer-specific analyses, there was evidence for increased risk of dementia for those with lung or colon cancer albeit with wide confidence intervals, but not for those with breast cancer. In contrast, beneficiaries with prostate cancer, particularly those diagnosed below the age of 50, appeared to have a lower risk of dementia than their cancer-free counterparts. Risk of death was higher among PWH with most cancers as compared those without. Notably, risk of death was substantially higher for PWH with lung cancer, but similar for PWH with and without prostate cancer. Thus, PWH with most cancers were more likely to die prior to development of dementia and thus likely to have attenuated risk differences of dementia between those with and without cancer. Collectively, these findings contribute to our understanding of the impact of cancer diagnoses on dementia risk among PWH while also highlighting the heterogeneity across cancer types and age at diagnosis.

Our finding that beneficiaries diagnosed with an ADC have an increased risk of dementia is consistent with prior literature that has reported an increased risk of dementia among those with AIDS and an increased risk of HIV-associated neurocognitive disorder (HAND) among those with poorly controlled HIV.^[1, 3]^ To address the influence of HIV care, we accounted for the use of ART, which we previously reported was less than 50% in this population^[12]^. However, we were unable to assess differences in viral suppression and CD4 count, which may influence cancer and dementia incidence. Of note, a larger proportion of ADCs in our study population occurred earlier in calendar time than for the other cancer types assessed, a pattern consistent with the declining incidence of ADCs reported in national trends.^[13]^ Nonetheless, these findings highlight the importance of access to and maintenance of high-quality HIV care to improve HIV and related health outcomes among PWH.

We found a modestly increased risk of dementia after diagnosis of any NADC, and this association was stronger among beneficiaries diagnosed with cancer below the age of 50. To our knowledge, no prior study has evaluated the association between a recent diagnosis of an NADC and dementia among PWH. Findings were similar in cancer-specific analyses for lung and colon cancer, although confidence intervals had decreased precision. While the mechanisms underlying these associations are unknown, we previously reported that cancer treatment, in particular chemotherapy and/or radiotherapy, was associated with significantly reduced CD4 count in adults with HIV ^[14]^. Low CD4 count has been associated with increased risk of HAND and dementia among people with HIV ^[15]^. More work is needed to examine the impact of cancer-specific treatments on HIV-specific outcomes and cognitive impairments.

Interestingly, a diagnosis of breast cancer or prostate cancer was not associated incidence of dementia. Indeed, men diagnosed with prostate cancer below the age of 50 had a lower incidence of dementia than their cancer-free counterparts. During the study period, national screening rates for breast and prostate cancer were high^[16, 17]^. We previously reported that PSA screening in Medicaid was slightly lower among Medicaid beneficiaries as compared to the general population, but comparable among men with and without HIV ^[18]^. Breast and prostate cancer screening protocols have been revised due to concerns they were associated with over detection of clinically insignificant cancer ^[19, 20]^. Although we were unable to assess cancers by stage, it is possible at least a sub-set of these cancers were over detected. Screen detected cancers may be a surrogate for more interaction with the healthcare system and/or less intensive cancer treatment, which may influence the diagnosis of dementia. In addition to treatment type, future studies should examine whether dementia risk is comparable among those diagnosed with local and later stage disease.

We evaluated the cumulative incidence of dementia among PWH with and without cancer who were enrolled in Medicaid in 14 states over a 15-year period. Approximately 40% of PLWH in the US are covered by Medicaid ^[9]^. Thus, our study population reflects a sizable proportion of PLWH in the US. We used an ICD-based definition of cancer,^10^ which we previously used to demonstrate that patterns of cancer incidence among PWH in Medicaid were comparable to those reported in other large, US-based studies ^[21-23]^.

We also used an ICD-based definition of dementia with strong positive predictive value among PWH ≥50 in large US healthcare system ^[24]^. Because HAND does not have a dedicated diagnosis code, we cannot rule out HAND, particularly among beneficiaries <50. In our study, PWH with cancer were similar to PWH without cancer with respect to demographic factors due to matching and to comorbidities and ART due to weighting. Nevertheless, differences in unmeasured factors, such as smoking status, could influence both cancer and dementia incidence. We conducted sensitivity analyses to compare PWH with cancer to cancer-free controls with conditions that could influence both cancer and dementia, including smoking associated conditions for lung cancer and metabolic conditions for colon cancer. We cannot rule out the potential for detection bias among PWH with cancer, though our sensitivity analyses may minimize this concern for lung and colon cancers. Prior work has highlighted potential sources of bias in analyses of the association between cancer and dementia in the general population.^[7, 8]^ Our study design and statistical analysis addressed several of these biases. For example, by assessing incident dementia prospectively from time of cancer diagnosis, we avoided the potential for survival bias incurred by restricting to those with dementia and avoided adjusting for factors affected by cancer diagnosis or treatment. We also explicitly used methods that allowed us to assess the association between cancer and dementia, while allowing for the competing event of death to occur.

We evaluated the incidence of dementia within 5 years of cancer diagnosis among PWH. We found that dementia risk increased among PWH with ADCs, any NADC, lung and colon cancer, but not breast or prostate cancer. Risk of death was higher among PWH with cancer as compared to PWH without cancer, however, there was substantial heterogeneity in risk differences by cancer type. The association between cancer and dementia was stronger among beneficiaries diagnosed below the age of 50; though this was largely due to increased dementia incidence among older beneficiaries without cancer. Collectively, these findings highlight the importance of consistent access to high quality HIV-specific and other healthcare. Given NADCs are increasing among PWH, more work is needed to understand the role of cancer stage and treatment type on risk of dementia among PWH.

## Data Availability

The data that support the findings of this study are under the authority of the Centers for Medicare & Medicaid Services (CMS) and administered by ResDAC. Investigators may reuse these data if they independently meet CMS requirements and obtain both CMS reuse approval and permission from the study’s NIH program officer.

