## Supplemental Data for "Incidence of dementia after a recent cancer diagnosis among people with HIV"

**Supplemental Methods**

We matched beneficiaries by cancer status for incident diagnoses of ADCs, NADCs and lung, breast, colon, and prostate cancers. We used incidence density matching with replacement to match beneficiaries with cancer to beneficiaries without cancer at the date of cancer diagnosis. Beneficiaries with cancer diagnosis after $b_0$ (the exposed) were sorted ascendingly by date of diagnosis (Figure S1). Using the sorted data, the matching pool for exposed beneficiary *i* was composed of all the unexposed at the time of cancer diagnosis $t_i$ (index date), regardless of future exposure. Any unexposed in the matching pool who had a dementia diagnosis, died, turned 65 years old, or ended Medicaid enrollment before $t_i$ was removed. The beneficiaries in the matching pool were matched to the exposed by age at $b_0$ +/- 2 years, sex (for NADC, ADC, lung, and colon cancers), race/ethnicity, United States (US) state, and year at $b_0$ +/- 2 years. Each exposed and all its matches formed a cluster. For each cluster *i*, the index date, $t_i$, was set as the new analytic baseline. Matches with any cancer or dementia diagnosis prior to $t_i$ were excluded. Due to the high number of NADC and ADC diagnoses, one match was selected randomly for each exposed. ART initiation and adherence were then identified per included beneficiary.

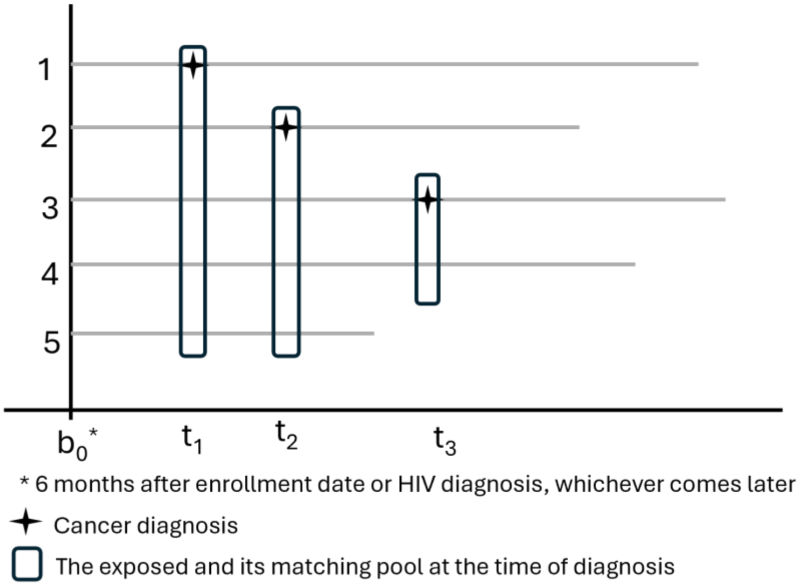

Figure S1. Diagram illustrating the steps on how the matching pool for the exposed group were obtained.

We then estimated the crude and weighted cumulative incidence functions (CIFs) of dementia among those with and without cancer using the Aalen-Johansen estimator to account for the competing risk of death, employing robust variance estimation to account for matching. Weighted CIFs were estimated using inverse probability weighting; exposure (cancer) weights and informative censoring weights were estimated and combined Weighted CIFs were estimated using inverse probability weighting; exposure (cancer) weights and informative censoring weights were estimated and combined. The exposure weight included age at enrollment (specified using natural cubic splines), MI, CHF, PVD, CVD, and diabetes status at baseline, number of Charlson comorbidities at baseline (0, 1 ≥2), and any ART use (weight 1) or ART adherence (weight 2). The censoring weight per beneficiary per record was computed as a ratio of two probabilities obtained using pooled logistic regression: 1) probability of not dropping out with time-updated age (specified as natural cubic splines) as the only covariate, and 2) the probability of not dropping out with the covariates time-updated age, exposure status, and time-varying Charlson comorbidity category (0, 1, ≥2) (within current interval, lag 3 months, and lag 6 months). We estimated the cumulative censoring weight per beneficiary per record by multiplying the censoring weights from the first through the last record and combined the exposure and censoring weights by multiplying the exposure weight to each record-specific cumulative censoring weight. The distributions of the combined weights were assessed and truncated at the 97.5$^{th}$ percentile. We calculated the crude and weighted risks of dementia and death at 1, 2, and 5 years after analytic baseline for beneficiaries with and without cancer and the corresponding risk differences (RD). The 95% confidence intervals (CI) of the risks and RDs were estimated using the 2.5th and 97.5th percentile of the point estimates from 500 bootstrap resamples. Analyses were also stratified by cancer type and age at index date (18-49; 50-64 years).

**Supplemental tables and figures**

Table S1. List of ICD-9 codes used in the study.

| Variable | ICD-9 Code[a] |
|---|---|
| **HIV** | 042-044, 079.53, 795.71, V08 |
| **Cancers** | |
| Lung | 162.2-162.5, 162.8, 162.9 |
| Colon | 153.X |
| Breast | 174.X, 175.X |
| Prostate | 185.X |
| ADC | 176.X, 180.X, 200.X, 202.X |
| NADC | 140.X-175.X, 177.X-179.X, 181.X-195.X, 199.X, 201.X, 203.X-208.X |
| Any cancer | 140.X-195.X, 199.X-208.X |
| **Dementia** | 046.19, 290.4x, 294.0, 294.1x, 294.2x, 294.8, 294.9, 331.0, 331.1x, 331.5, 331.6, 331.7, 331.8x, 331.82, 438.0, 781.8 |
| **Charlson comorbidities** | |
| Myocardial infarction | 410.x, 412.x |
| Congestive heart failure | 398.91, 402.01, 402.11, 402.91, 404.01, 404.03, 404.11, 404.13, 404.91, 404.93, 425.4-425.9, 428.x |
| Peripheral vascular disease | 093.0, 437.3, 440.x, 441.x, 443.1-443.9, 447.1, 557.1, 557.9, V43.4 |
| Cerebrovascular disease | 362.34, 430.x-438.x |
| Chronic pulmonary disease | 416.8, 416.9, 490.x-505.x, 506.4, 508.1, 508.8 |
| Rheumatic disease | 446.5, 710.0-710.4, 714.0-714.2, 714.8, 725.x |
| Peptic ulcer disease | 531.x-534.x |
| Mild liver disease | 070.22, 070.23, 070.32, 070.33, 070.44, 070.54, 070.6, 070.9, 570.x, 571.x, 573.3, 573.4, 573.8, 573.9, V42.7 |
| Diabetes without chronic complication | 250.0-250.3, 250.8, 250.9 |
| Diabetes with chronic complication | 250.4-250.7 |
| Hemiplegia or paraplegia | 334.1, 342.x, 343.x, 344.0-344.6, 344.9 |
| Renal disease | 403.01, 403.11, 403.91, 404.02, 404.03, 404.12, 404.13, 404.92, 404.93, 582.x, 583.0-583.7, 585.x, 586.x, 588.0, V42.0, V45.1, V56.x |
| Moderate or severe liver disease | 456.0-456.2, 572.2-572.8 |

[a] A code ending with ".X" indicates a wildcard. Any number could appear after the decimal place.

A. Crude estimates

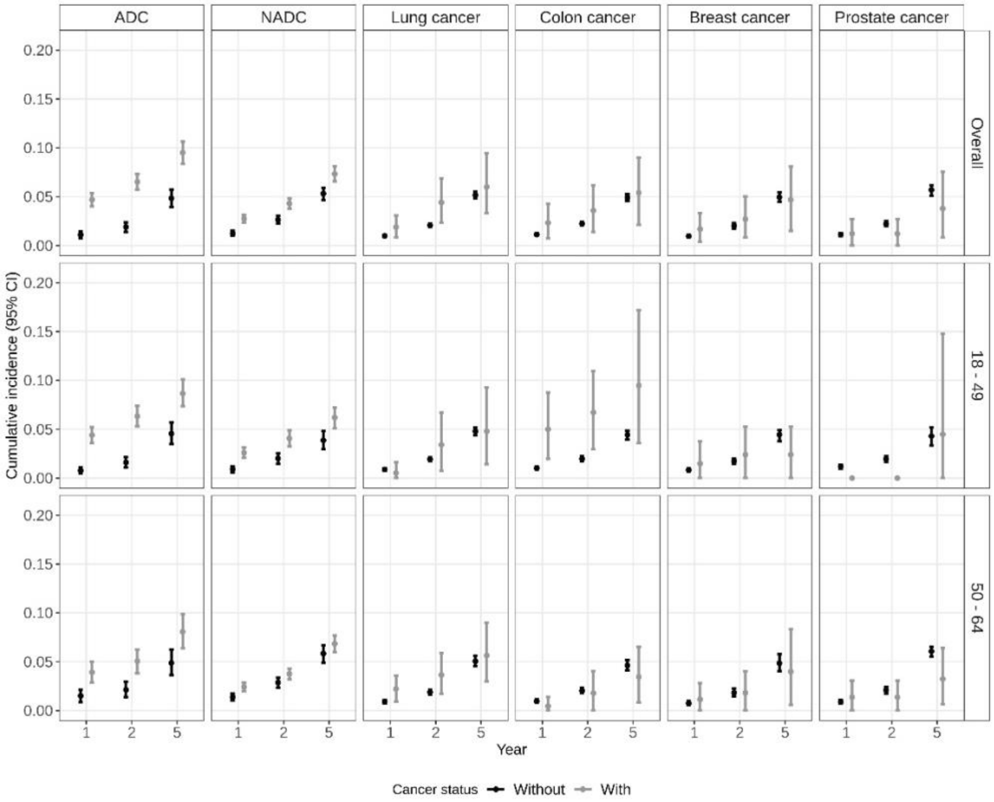

B. Weighted estimates (weight 1)

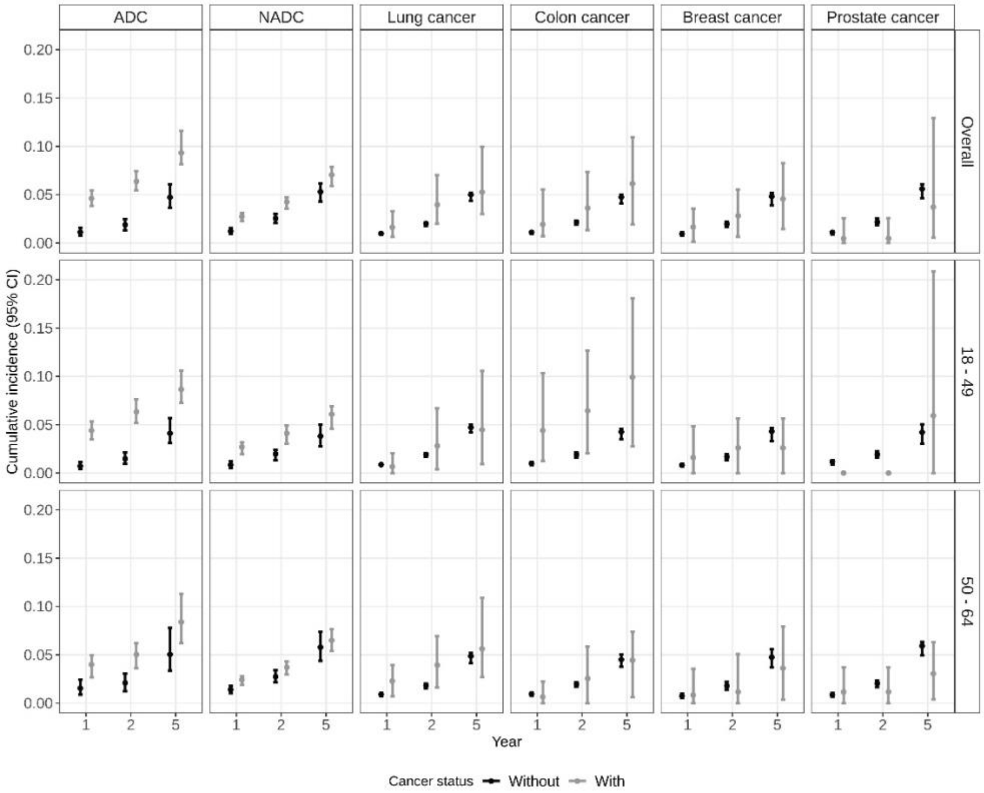

Figure S2. A. Crude and B. weighted cumulative incidence (risk%) of dementia at years 1, 2, and 5 after baseline among matched Medicaid beneficiaries with HIV by cancer status, overall and stratified by age.

A. Crude estimates

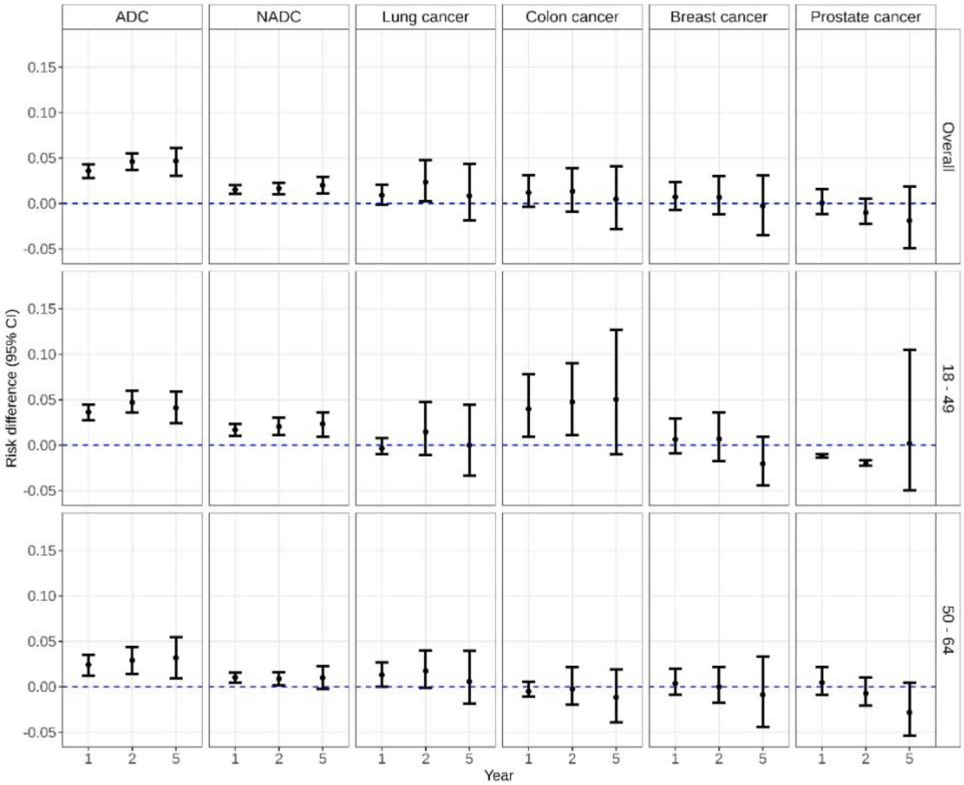

B. Weighted estimates (weight 1)

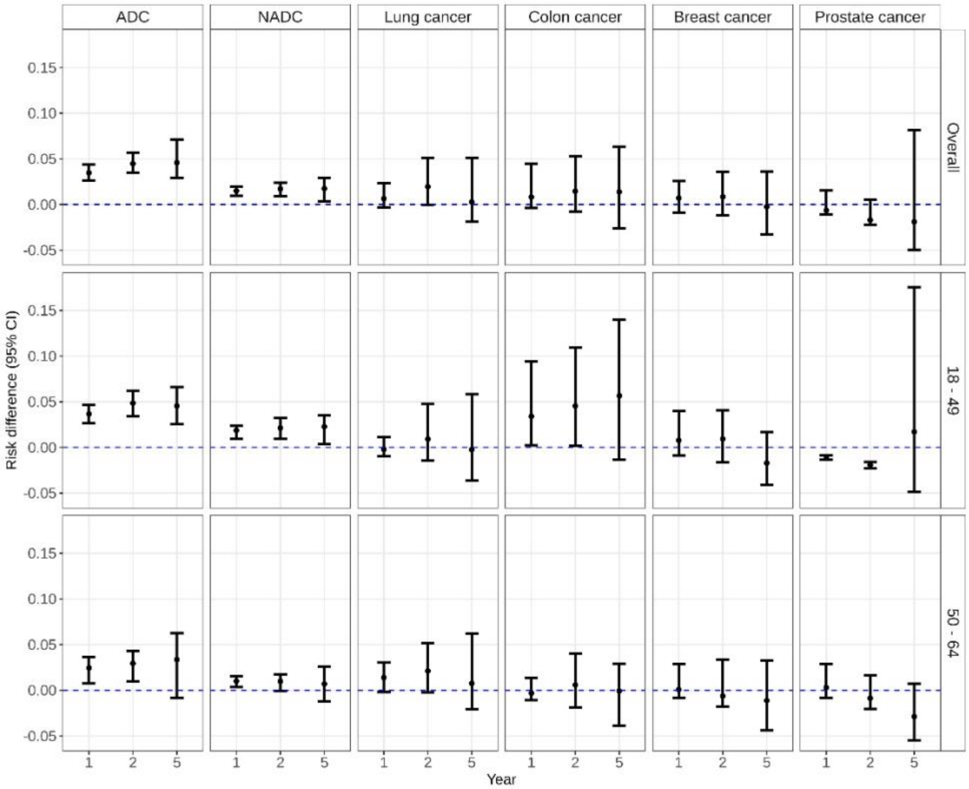

Figure S3. A. Crude and B. weighted risk difference of dementia of those with and without cancer at years 1, 2, and 5 after baseline among matched Medicaid beneficiaries with HIV by cancer status, overall and stratified by age.

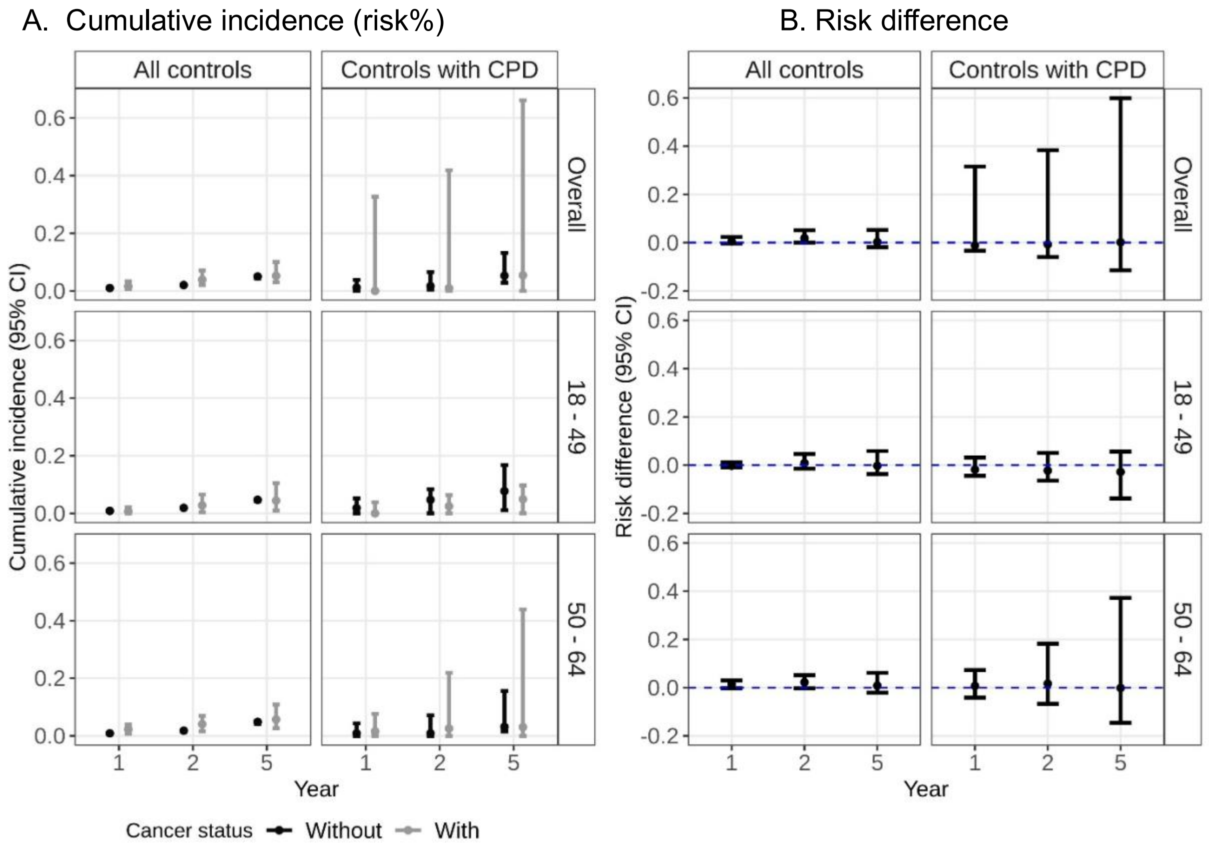

Figure S4. A. Weighted cumulative incidence (risk%) and B. weighted risk difference of dementia at years 1, 2, and 5 after baseline comparing Medicaid beneficiaries with lung cancer and their matched controls with chronic pulmonary disease (CPD), overall and stratified by age.

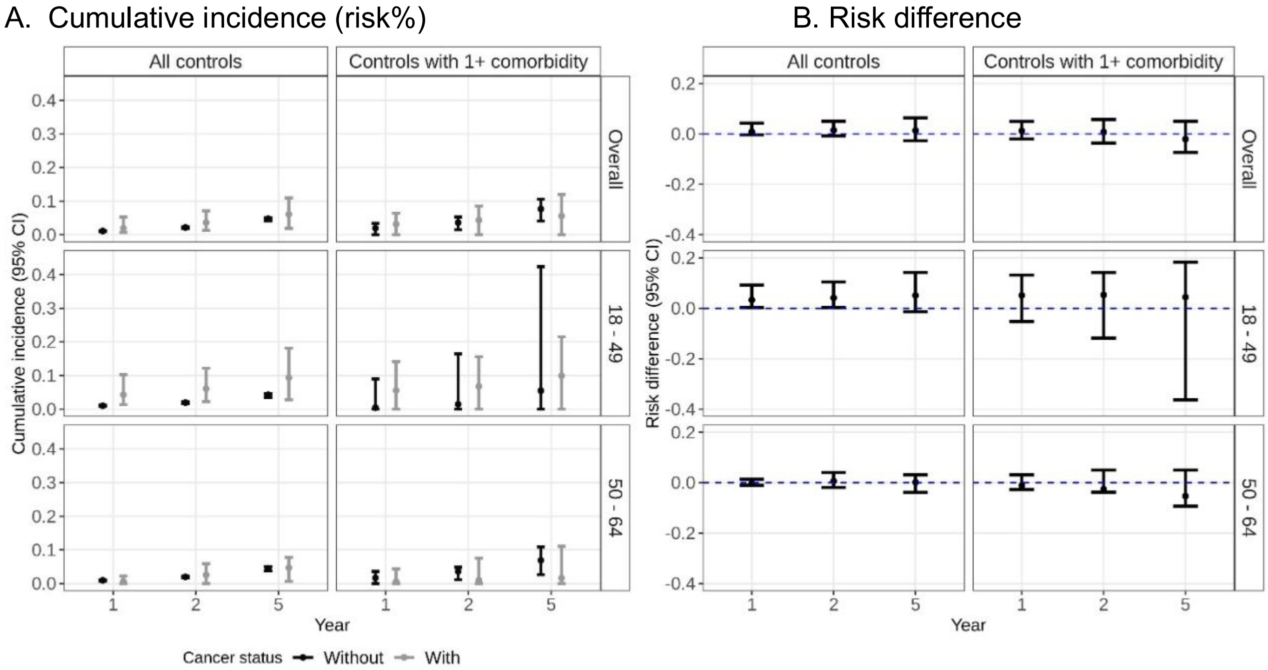

Figure S5. A. Weighted cumulative incidence (risk%) and B. weighted risk difference of dementia at years 1, 2, and 5 after baseline comparing Medicaid beneficiaries with colon cancer and their matched controls with at least 1 comorbidity, overall and stratified by age.

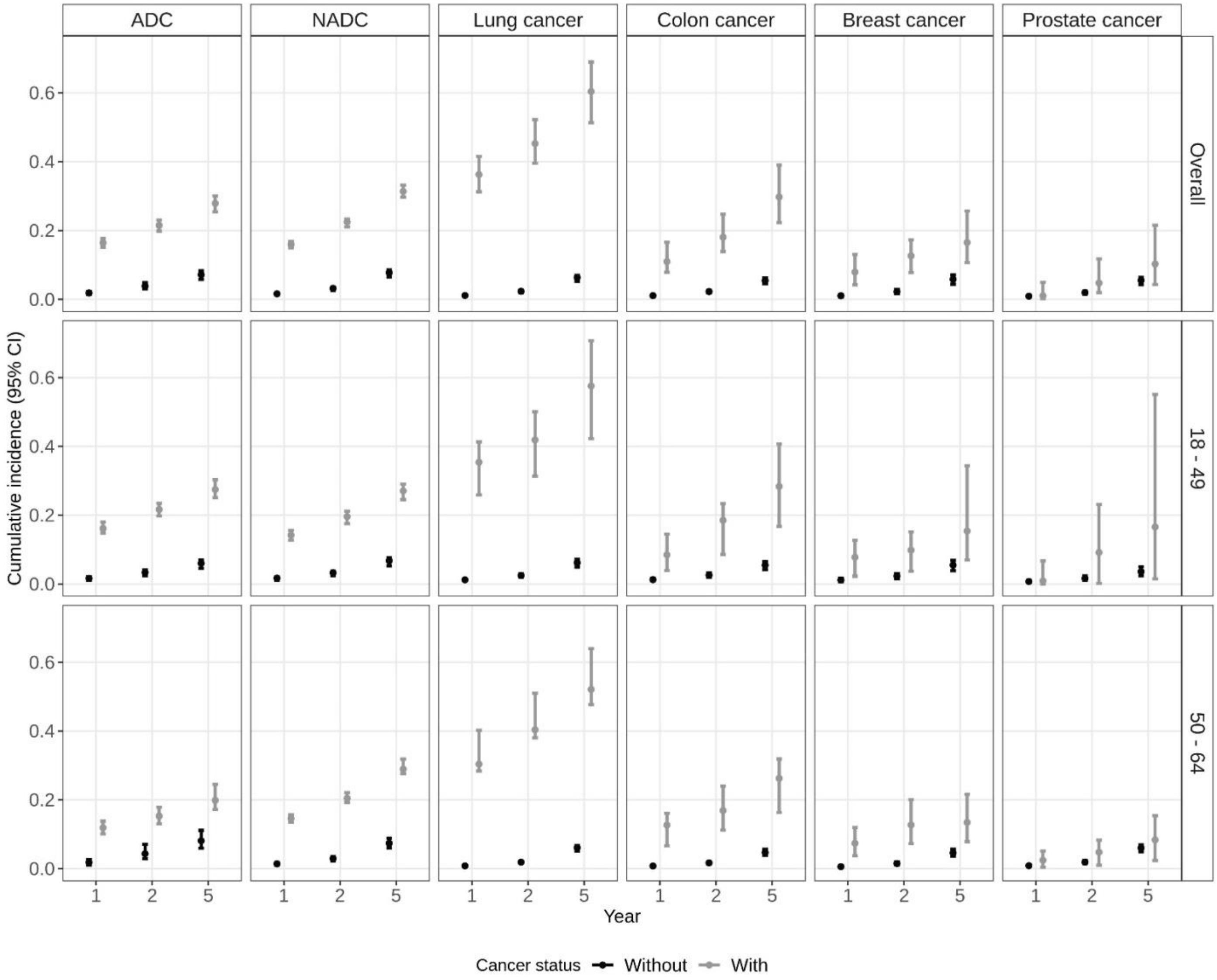

Figure S6. Weighted cumulative incidence (risk%) of death at years 1, 2, and 5 after baseline among matched Medicaid beneficiaries with HIV by cancer status, overall and stratified by age.

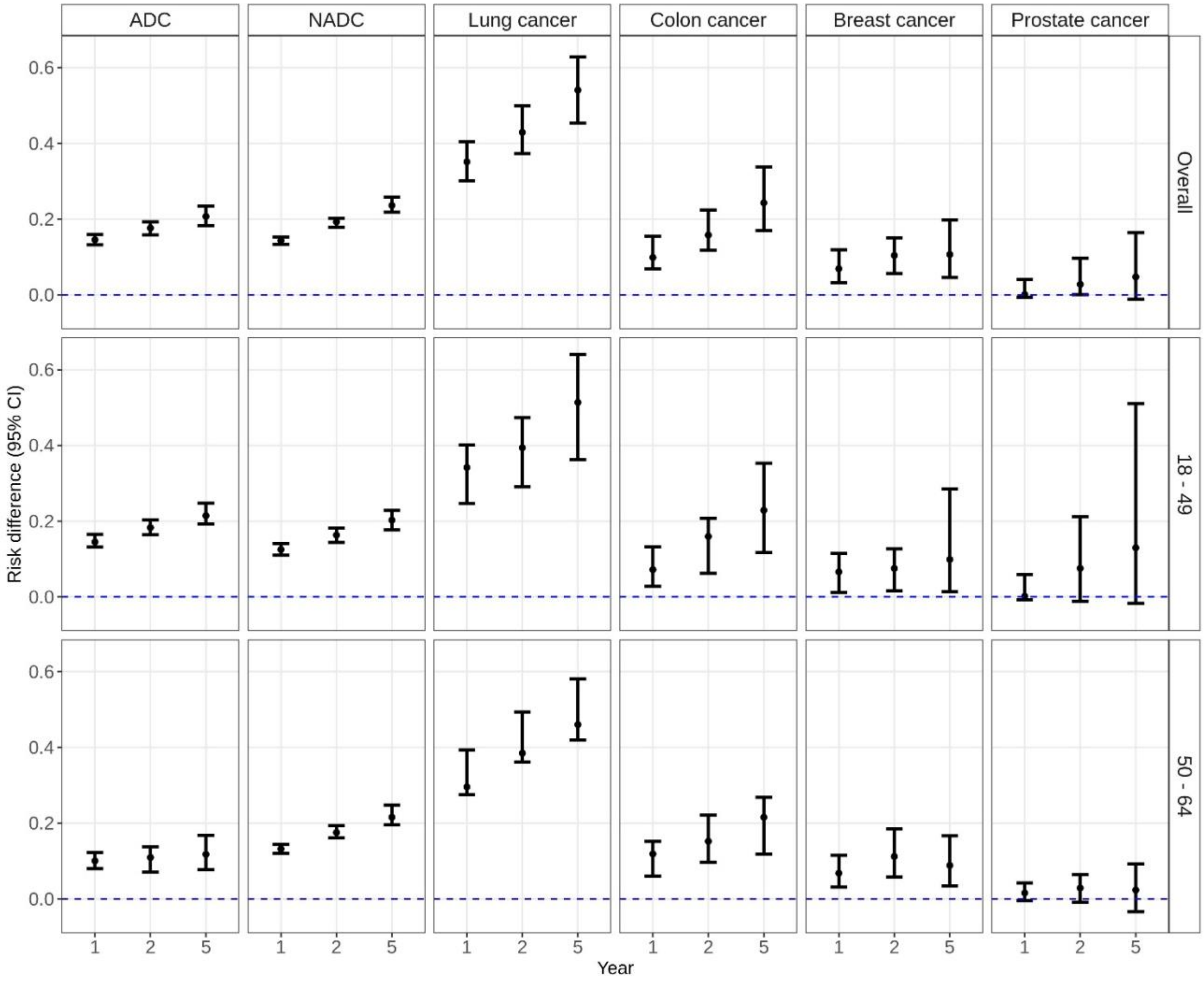

Figure S7. Weighted risk difference of death at years 1, 2, and 5 after baseline of those with and without cancer among matched Medicaid beneficiaries with HIV by cancer status, overall and stratified by age.

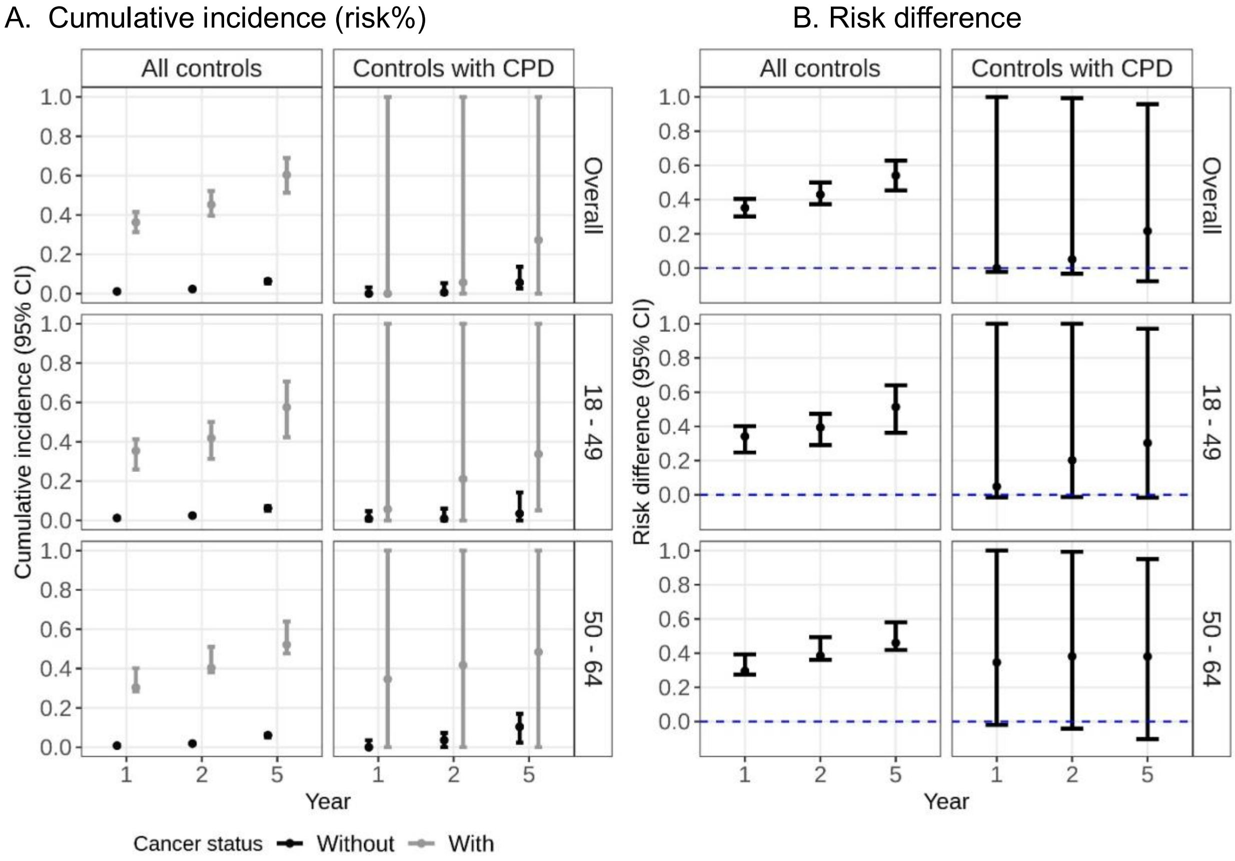

Figure S8. A. Weighted cumulative incidence (risk%) and B. weighted risk difference of death at years 1, 2, and 5 after baseline comparing Medicaid beneficiaries with lung cancer and their matched controls (all controls – 1st column of each panel, controls with chronic pulmonary disease (CPD) – 2nd column of each panel), overall and stratified by age.

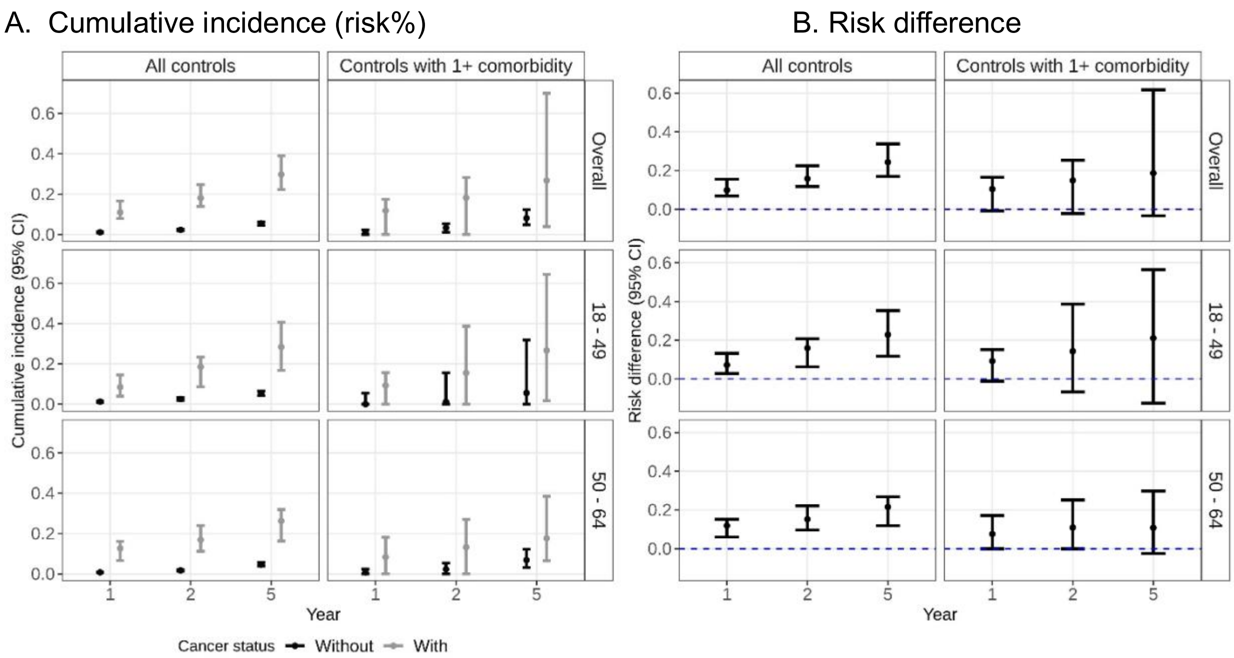

Figure S9. A. Weighted cumulative incidence (risk%) and B. weighted risk difference of death at years 1, 2, and 5 after baseline comparing Medicaid beneficiaries with colon cancer and their matched controls (all controls – 1st column of each panel, controls with at least 1 comorbidity – 2nd column of each panel), overall and stratified by age.
